# Analysis of IL-4 Levels, IL-1α, Cortisol Hormone, and Blood Cell Number in Parkinson’s Disease Patients Undergoing Dance-based Movement Therapy: A Preliminary Study

**DOI:** 10.1101/2024.09.21.24314141

**Authors:** Karen Adriana Pastana Marques, Gabriel Mesquita da Conceição Bahia, Juliana dos Santos Duarte, Denise da Silva Pinto, Arnaldo Jorge Martins Filho, Evander de Jesus Oliveira Batista, Anderson Manoel Herculano Oliveira da Silva, Karen Renata Herculano Matos Oliveira, Lane Viana Krejcova, Carlomagno Pacheco Bahia

## Abstract

**Introduction:** The interplay between neuroinflammation and Parkinson’s disease (PD) is still unclear, although several studies suggest that neuroinflammation plays an important role in the pathophysiological pathways of neurodegenerative disorders. Dance-based movement therapy has been studied as an adjuvant treatment for PD symptoms by modulating some inflammatory interleukins and improving the overall effect of pharmacological treatments.

**Objective:** To evaluate the levels of Interleukin 4, Interleukin 1α, cortisol hormone, and blood cell total numbers in PD subjects undergoing dance-based movement therapy.

**Methods:** Twenty-five participants were selected for this study. Seventeen subjects diagnosed with PD were divided into two groups: PD group (n=8) and PD+dance group (n=9). Another group was composed of age-matched healthy controls (n=8). The dance-based movement therapy was performed twice a week for 50-60 minutes/session over six months.

**Results:** The serum IL-4 levels were significantly lower in the PD group when compared to the healthy group, while the PD+Dance group showed similar levels to the healthy group. The levels of IL-1α, cortisol, and blood cell count showed no statistical differences between groups.

**Conclusion:** Our results suggest that regular practice of dance-based movement therapy may induce a neuroprotective effect by modulating the levels of IL-4 in people with PD similarly to healthy participants.

**Trial registration:** The study was registered in the Brazilian registry of clinical trials: RBR-3bhbrb5.

## Introduction

The interplay between inflammation and Parkinson’s disease (PD) is still unclear, although inflammation plays an important role in neurodegenerative disorders[1] and may help promote neuronal death and involve blood cell abnormalities[2].

The effects of physical exercise on motor and non-motor symptoms in people with PD effectively modulate the inflammatory process[3], promoting neuronal survival by increasing the production of neurotrophic factors, releasing neurotransmitters, and improving neurogenesis, angiogenesis, modulating inflammatory cytokines, and cortisol hormone expression[3,4].

Thus, therapy based on dance movements may be a physical activity alternative to improve PD treatment [5], once dance involves motor skills such as motion planning, rhythm, motor memory, proprioception, imitation[5–7], emotional aspects, and visuospatial information, in addition to increasing body balance, integrating brain areas, decreasing cortisol hormone, and improving neuroplasticity [5,6,8,9].

To this date, no study has analyzed the serum levels of inflammatory cytokines, the stress hormone (cortisol), and the blood cell number in people with PD undergoing dance-based movement therapy. Therefore, the present study evaluated the serum levels of Interleukin 4 (IL-4), Interleukin 1α (IL-1α), cortisol hormone, and total blood cell numbers in people with Parkinson’s disease undergoing physical activity based on dance movements.

## Methods

### Study design and Participants

We performed a longitudinal study aiming to analyze the effects of physical activity based on dance movements using the “Baila Parkinson” method as a complementary therapy for Parkinson’s disease [10].

We submitted this study to the ethical committee of the Federal University of Pará (CEP: 3.965.311/ CAAE: 27811119.4.0000.0018) and registered it in the Brazilian Registry of Clinical Trials (ReBEC RBR-3bhbrb5, UTN code: U1111-1275-6679). The research was developed from July to December 2019, the participants were recruited through social media announcements, the follow-up was until march 2020, and the inclusion criteria were: 1) a diagnosis of PD according to the UK Parkinson’s Disease Society Brain Bank (UKPDSBB); 2) Hoehn and Yahr stages I to III under pharmacological treatment for at least 3 years; and 3) the ability to walk independently. Exclusion criteria: 1) presence of other neurologic or neuropsychiatric conditions; 2) comorbid inflammatory disease; or 3) severe cardiopulmonary disease. This study was conducted in accordance with the Declaration of the World Medical Association and all participants provided written informed consent.

The study included 25 subjects. Seventeen (n=17) subjects with Parkinson’s disease, and eight (n=8) healthy controls (Healthy Group - HG) were selected. Demographic and clinical parameters were documented. Among the subjects with PD, eight received their PD medical treatment as usual (PD Group, n=8), whereas the other nine subjects with PD received medical treatment associated to dance therapy (PD+Dance, n=9). All PD subjects were submitted to evaluation of disease progression through the Unified Parkinson Disease Rating Scale (UPDRS), and fulfilled UKPDSBB clinical criteria for idiopathic PD (H&Y Scale 1,88 ± 0,33; UPDRS III 30,55 ± 13,60; UPDRS Total 48,77 ± 19,63).

### Intervention: Dance-based movement therapy

The dance-based movement therapy was performed twice a week, 50-60 minutes per session for six months following a protocol based on the “Baila Parkinson” method [10].

### Data Collection and Statistical Analysis

We collected the peripheral blood of all participants (10 mL through peripheral vein puncture; at 8-9 am after 8h of fasting) and centrifuged it at 3500 g for 10 min. The serum was removed and stored at –80 °C (Sanyo VIP Plus -80°C Freezer) until analysis.

The levels of IL-4, IL-1α, and cortisol were measured using the Enzyme-Linked Immunosorbent Assay - ELISA (Invitrogen®) method. The blood cell number was calculated using impedance and colorimetric measurements (Medical Laboratory Equipment Blood Test CBC). The results were analyzed using GraphPad Prism® 8.0 Software. We tested the data for normality (Kolmogorov-Smirnov) and homogeneity (Levine test). The data were analyzed using a one-way analysis of variance (ANOVA) corrected by the Tukey intergroup post-test or Kruskal-Wallis test for non-parametric data. Statistical significance was p<0.05, and all data are presented as mean ± standard deviation (M±SD).

## Results

The present study aimed to the serum levels of Interleukin 4 (IL-4), Interleukin 1α (IL-1α), cortisol hormone, and total blood cell numbers in people with Parkinson’s disease undergoing physical activity based on dance movements. Peripheral blood was collected once and IL-1α, IL-4, cortisol levels and blood cell count were measured.

Twenty-five individuals enrolled in this study which eight were healthy and seventeen were with idiopathic PD. Nine individuals with idiopathic PD were selected to attend the therapeutic sessions (Fig 1). were selected to attend the therapeutic sessions (Fig 1). The demographic and clinical parameters of the three subjects’ groups are shown in Table 1. There were no statistically significant differences in age or gender among the three groups.

**Table 1.** Demographic and clinical features of the three groups.

| Demographic and Clinical Features | Parkinson's Group with Dance (PD + Dance)<br>N=9 | Parkinson's Without Dance Group (PD)<br>N=8 | Healthy Group (HG)<br>N=8 |
| --- | --- | --- | --- |
| Sex |  |  |  |
| Male | 4 | 4 | 3 |
| Female | 5 | 4 | 5 |
| Age (years) | 63,22 ± 9,40 | 67,25 ± 13,41 | 59,12 ± 10,73 |
| Time since Diagnosis (Years) | 4,55 ± 2,50 | 7,22 ± 5,80 | - |
| H&Y Scale | 1,88 ± 0,33 | - | - |
| UPDRS III | 30,55 ± 13,60 | - | - |
| UPDRS Total | 48,77 ± 19,63 | - | - |

UPDRS: Unified Parkinson's disease rating scale; H&Y Scale: Hoehn and Yahr Scale

**Figure 1.**
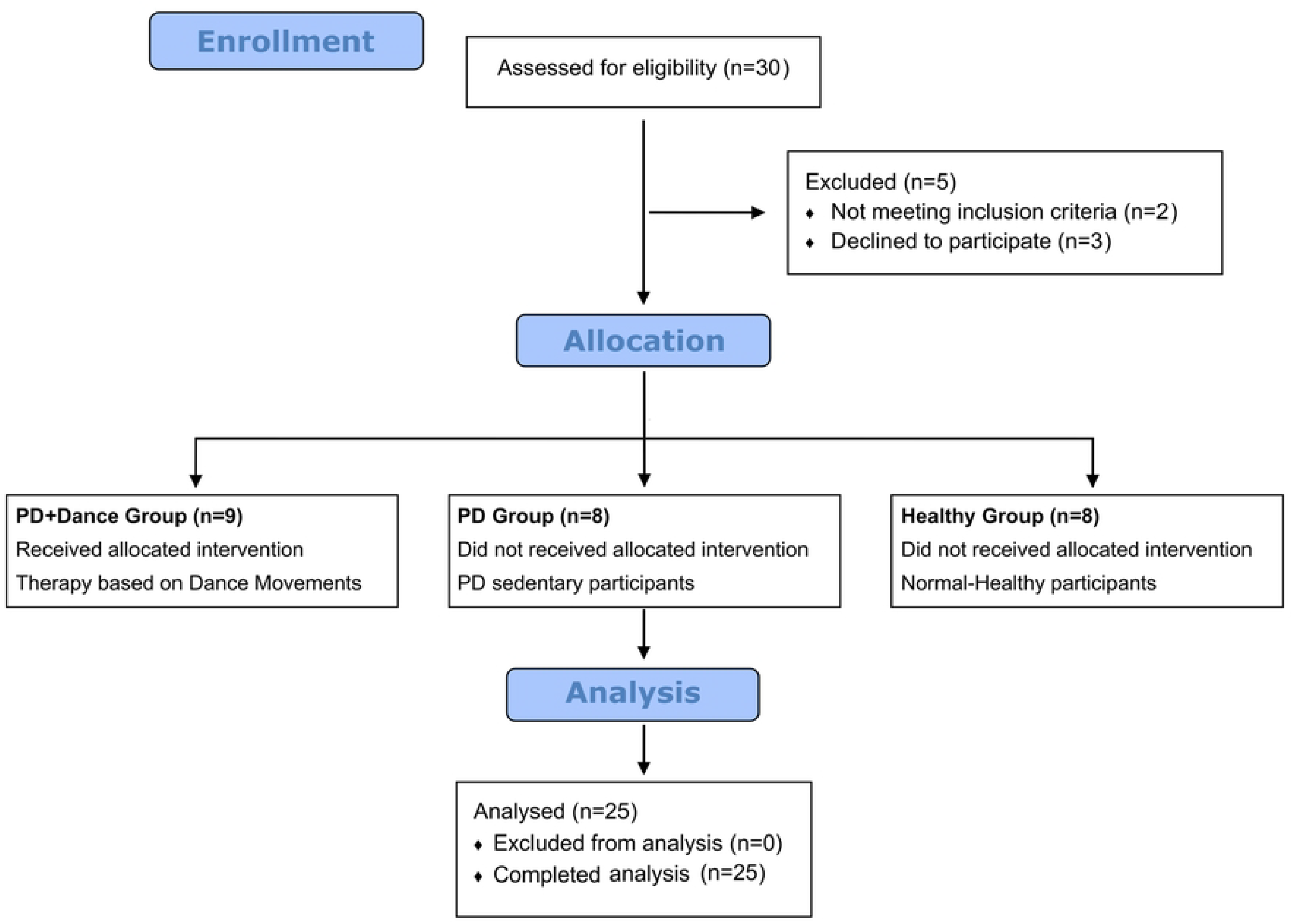
Flowchart of the participant’s recruitment for the study.

### Pro and Anti-Inflammatory Cytokine Levels

Serum blood levels of IL-4 in the PD group were significantly lower when compared to the healthy group (HG) while the PD+Dance group had IL-4 serum levels similar to the healthy group (HG: 0.37± 0.05; PD: 0.1280±0.02; PD+Dance: 0.2300± 0.08; p=0,0079). Regarding the levels of IL-1α, there was no statistically significant difference between the groups, but the concentration of IL-1α in the PD+Dance subjects was lower in relation to the PD group (HG: 0.008±0.004; PD group: 0.022±0.02; PD+Dance group: 0.01±0.01; p=0,5051), see Figure 2 A and B.

**Figure 2.**
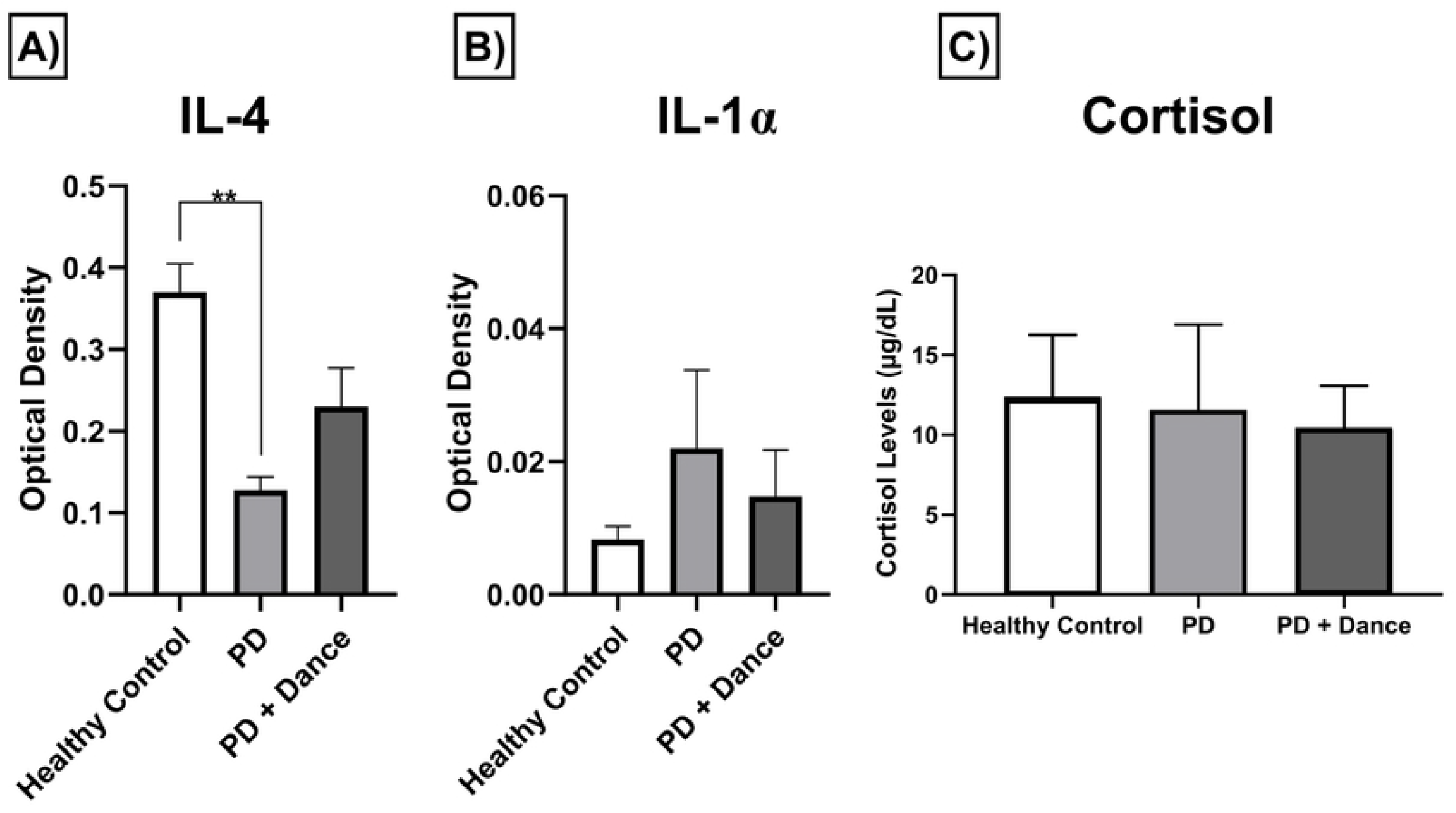
The blood levels of IL-4, IL-1α, and cortisol hormone analysis (µg/dL). Statistical difference assessed using one-way ANOVA test (p < 0.05). A) Serum levels of interleukin 4: HG 0.37 (± 0.05); PD 0.1280(± 0.02); PD + Dance 0.2300 (± 0.08; p=0,0079). B) Serum levels of interleukin 1α: HG 0.008 (± 0.004); PD 0.022 (± 0.02) and PD + Dance 0.01(± 0.01; p=0,5051). C) Serum levels of cortisol: HG 12.40±3.86; PD Group 11.57±5.31; PD + Dance Group 10.32± 2.75; ANOVA p=0,56.

### Serum Cortisol Levels

The serum blood level of cortisol hormone did not show statistically significant differences among the three groups analyzed (HG: 12.40±3.86; PD Group: 11.57±5.31; PD + Dance Group: 10.32± 2.75; ANOVA p=0,56), see Figure 2C.

### Blood Cells Count

The blood cell counts did not show statistical differences among the three groups (p>0.05), see Table 2. However, two participants of the PD group that underwent dance-based movement therapy (PD+Dance) had changes in red cell numbers, which may indicate anemia and positive microcytosis (see Table 2). There was also one case with erythrocytosis, one with hypochromic, another with thrombocytopenia, and the last with atypical lymphocytes (Table S1).

**Table 2.** Blood cell total numbers of the three groups (Data shown as mean ± standard deviation and one-way ANOVA test).

| Blood Cell Count | Parkinson's Group with Dance (PD + Dance)<br>N=9 | Parkinson's Without Dance Group (PD)<br>N=8 | Healthy Group (HG)<br>N=8 | p Value |
| --- | --- | --- | --- | --- |
| Red Blood Cells($10^6/\text{mm}^3$) | 4848888,88 ± 589246,22 | 4878750 ± 817879,44 | 5120000 ± 571564,26 | >0,05 |
| Hemoglobin (g/dl) | 14,28 ± 2,16 | 14,60 ± 2,99 | 14,77 ± 1,40 | >0,05 |
| Hematocrit (%) | 42,98 ± 6,24 | 44,48 ± 8,67 | 44,37 ± 4,13 | >0,05 |
| MCV ($\mu\text{m}^3$) | 88,56 ± 6,49 | 90,92 ± 4,07 | 87,03 ± 3,33 | >0,05 |
| MCH (pg) | 29,42 ± 2,42 | 29,81 ± 1,97 | 28,96 ± 1,32 | >0,05 |
| MCHC (g/dl) | 33,21 ± 0,72 | 32,78 ± 0,96 | 33,77 ± 1,41 | >0,05 |
| White BloodCells ($10^3/\text{mm}^3$) | 6503,33 ± 1320,70 | 7070 ± 1417,58 | 6343,75 ± 1079,53 | >0,05 |
| Lymphocytes (p/$\text{mm}^3$) | 2033,22 ± 646,03 | 2173,25 ± 461,21 | 2333,50 ± 567,02 | >0,05 |
| Monocytes (p/$\text{mm}^3$) | 607204,23 ± 136,84 | 400,50 ± 87,34 | 438,75 ± 130,26 | >0,05 |
| Platelets ($10^3/\text{mm}^3$) | 233111,11 ± 36285,13 | 205250 ± 46092,29 | 222875 ± 32148,92 | >0,05 |
| MPV ($\mu\text{m}^3$) | 8,48 ± 1,24 | 10,93 ± 3,82 | 8,81 ± 0,96 | >0,05 |

MCV: Mean Corpuscular Volume; MCH: Mean Corpuscular Hemoglobin; MCHC: Mean Corpuscular Hemoglobin Concentration; MPV: Mean Platelet Volume.

## Discussion

The participants in the PD group presented lower blood levels of IL-4 when compared to the HG group. The PD+Dance group had IL-4 concentration levels similar to those of the HG. This finding is relevant given the importance of IL-4 as a regulator for the inflammation process and its relationship with normal neuronal function which is damaged in neurodegenerative diseases[11]. Once physical activity may regulate different kinds of inflammatory cytokines, inhibiting neuroinflammatory responses via the upregulation of interleukins [12], our results suggest that dance-based movement therapy modulated neuroprotective mechanisms across the increasing IL-4 blood serum concentrations in the PD+Dance group (Figure 2A). However, it is necessary to evaluate the cellular and molecular mechanisms involved in the effects of regular therapy based on dance movements.

There was no difference among the three groups for IL-1α serum blood levels. However, the IL-1α blood serum levels of the PD+Dance group showed lower absolute values compared to the PD group (Figure 2B). Once the cell death is directly related to the regulation of IL-1α processing and release, conceivably the neurodegenerative and chronic character of Parkinson’s disease must be inducing cell death across the constant release of IL-1α[1,12].

There was no difference among the three participants’ groups regarding serum cortisol blood levels (Figure 2C). Although our results were not statistically significant, our work demonstrated that therapy based on dance movements was able to conserve cortisol levels in people with PD.

There was no statistically significant difference among the three experimental groups regarding the total number of blood cells. For the parameters of blood cell counting, see Table 2. However, the proportion of red blood cells and the platelet count of both the PD+Dance Group and the PD Group showed alterations in some cases, such as anemia and microcytosis (Table S1). The number of red blood cells, leukocytes, and hematocrit values were not affected by the experimental conditions and were similar to a previous study that supports the association between low hemoglobin levels and PD [13] [14].

## Conclusion

Dance-based movement therapy reduced blood serum levels of IL-4, suggesting that its regular practice may modulate neuroprotective effective action to delay the progression of Parkinson’s disease.

## Data Availability

All relevant data are within the manuscript and its Supporting Information files.

## Acknowledgment

We thank the Health Science Institute and the Federal University of Para for their support and technical assistance.

